# Insights from a Ventilation-Aware Pandemic and Outbreak Risk Model (VAPOR)

**DOI:** 10.1101/2025.07.30.25332443

**Authors:** Natalie J. Wilson, Callandra Moore, Clara Eunyoung Lee, Ashleigh R. Tuite, David N. Fisman

## Abstract

Transmission of airborne pathogens in indoor spaces is strongly modulated by heterogeneity in ventilation. Understanding the role indoor air plays in pandemic risk is limited in part due to differing modeling approaches used in engineering and epidemiology. Here we present the VAPOR (Ventilation-Aware Pandemic and Outbreak Risk) model, a hybrid transmission framework that integrates Reed-Frost close-contact dynamics with Wells-Riley aerosol-mediated risk. Using a meta-population structure to simulate multi-patch environments (e.g., separate workplaces or schools), we explore how ventilation disparities shape epidemic potential. A fixed minority of individuals are modeled as “aerosolizers,” consistent with overdispersed real-world transmission patterns (e.g., SARS-CoV-2). Simulations reveal that both improving ventilation in high-risk patches and raising baseline ventilation across environments independently reduce risk. Parameter sweeps across air changes per hour (ACH, 2–12) demonstrate non-linear benefits with early saturation. These findings emphasize the need for targeted ventilation strategies and show how small-world effects amplify heterogeneity-driven transmission. VAPOR offers a framework for linking ventilation equity to epidemic control.

## Introduction

The COVID-19 pandemic served as a potent reminder of the damage airborne pathogens can cause (1–3). Airborne transmission is a dominant route for many respiratory infections, including both novel pathogens with pandemic potential and re-emerging ones tied to waning vaccine uptake (4–7). While both engineers and epidemiologists model infections spread through shared air, their approaches differ, shaped by disciplinary silos and divergent goals.

Engineers have historically approached the problem using models such as the Wells-Riley (WR) equation or (8), more recently, computational fluid dynamics (CFD), which estimate exposure based on ventilation, air mixing, and room geometry (9–11). Epidemiologists, by contrast, favor compartmental or stochastic models that simulate transmission through human contact networks, typically without explicit reference to air. These modeling traditions differ not just in assumptions and time scales, but in terminology: for example,’mixing’ refers to airflow in engineering (12) but to social interaction in epidemiology (13).

Despite these differences, there are conceptual bridges. Both Wells-Riley and the Reed-Frost model (a canonical small population stochastic outbreak model used in epidemiology) rely on Bernoulli trials to represent infection as a probabilistic event (8, 14). This shared logic allows us to reinterpret the Wells-Riley framework as a generator for the transmission parameter in Reed-Frost models, yielding a hybrid structure that captures multi-generational transmission with mechanistic attention to indoor air and ventilation.

A further challenge in modeling early transmission is the high heterogeneity in individual infectivity, often represented by a skewed negative binomial distribution with dispersion parameter *k* (15–17). For pathogens like SARS-CoV-2, a low *k* means that most cases generate few secondary infections, but a small subset cause superspreading events (18–21). Such events are especially influential early in an epidemic, where emergence or extinction depends on stochastic chains of transmission. These dynamics are driven both by biological and behavioral variability (e.g., host viral load, aerosol generating activities like singing) and by environmental risk factors such as poor ventilation (22–25).

Heterogeneity in host viral load may have been an important driver of the overdispersed *R*_0_ of SARS-CoV-2 seen early in the COVID-19 pandemic (18, 20, 21). Conceptually one might approach such heterogeneity by dividing populations into a majority of “non-aerosolizers” and a minority population of “aerosolizers”, whose infectivity depends on aerosol production and inadequate air exchange (4, 10, 22). Such a heterogeneous population can then be modeled using a hybrid approach that integrates Reed-Frost and Wells-Riley models into a common probabilistic architecture. We refer to this hybrid framework as the VAPOR (Ventilation-Aware Pandemic and Outbreak Risk) model, which can be used to examine disease emergence, extinction dynamics, and the effects of targeted interventions, ventilation improvements, and vaccination, all within a structure that makes indoor air explicit.

Our objective in this work is to apply this hybrid framework to gain insights into how heterogeneity in indoor ventilation influences the emergence and control of airborne epidemics. Specifically, we aim to determine how vulnerability to pathogen emergence is shaped by uneven air exchange, especially under conditions of overdispersed transmission, while providing a modeling framework that bridges engineering and epidemiological paradigms.

## Methods

### i. The Reed-Frost Model

The Reed-Frost model is a canonical discrete-time model for simulating the spread of directly transmitted infectious diseases in small, closed populations (14). The original model was developed in the 1920s using a physical model (a trough with coloured marbles) and was never published by Reed and Frost, but the model was described mathematically by Fine in 1977 (14). The model operates under a Bernoulli trial framework, assuming that infection spreads through “adequate contact” between susceptible and infectious individuals during discrete time steps (typically corresponding to one serial interval). For a population with *S_t_* susceptible and *C_t_* infectious individuals at time *t*, the probability that a given susceptible escapes infection is:

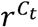

where *r*=1-*p* and *p* is the per-contact probability of infection. Thus, the expected number of new cases at time *t+1* is given by:

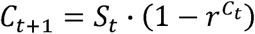

and the updated number of susceptibles is:

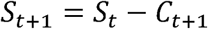

A stochastic formulation is:

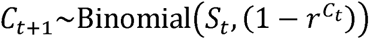

This latter version captures the possibility of stochastic extinction and variation in epidemic size due to chance, even under identical initial conditions.

### ii. The Wells-Riley Equation

The Wells-Riley equation is a canonical model in environmental engineering used to estimate the probability of infection due to airborne transmission in shared indoor spaces (8). It is derived from principles of mass balance and assumes well-mixed air, constant generation of infectious “quanta”, and steady-state ventilation. The Wells-Riley equation is based on a mass-balance approach, in which the number of infectious quanta (each quantum representing the number of infectious particles required to infect) in a space is determined by the rate at which they are generated by infectious individuals and the rate at which they are removed by ventilation.

Assuming well-mixed air and steady-state conditions, the model estimates the average concentration of quanta in air and computes infection probability as a function of the dose inhaled by a susceptible person over a defined period of exposure.

The probability that a susceptible person becomes infected during exposure to shared air is given by:

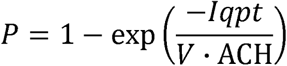

where *I* is the number of infectious individuals, *q* is the quanta generation rate per person, *p* is the pulmonary ventilation rate of susceptibles, *t* is the duration of exposure, and v·ACH is the clean air delivery rate, where ACH is air changes per hour and *V* is room volume. In the VAPOR model, this probability becomes the effective transmission rate for the subset of individuals who transmit via aerosol (the *C_b_*class as described below). Specifically, the Wells-Riley probability substitutes for the per-contact probability 1-*r^ct^* in the Reed-Frost model, thereby linking environmental parameters to the probability of transmission in a discrete-time, generational framework.

### iii. The VAPOR Model

We developed a hybrid infectious disease transmission model, the <u>V</u>entilation-<u>A</u>ware <u>P</u>andemic and <u>O</u>utbreak <u>R</u>isk (VAPOR) model, which synthesizes key elements of the Reed-Frost and Wells-Riley frameworks. The VAPOR model allows for recursive transmission over time via the Reed-Frost model while incorporating environmental determinants of airborne risk, such as ventilation, via the Wells-Riley equation. VAPOR assumes that individuals infected with an airborne pathogen can be divided into two classes:

a. *C_a_* (close contact) cases, who transmit infection through close proximity exposure, with transmission governed by a per-contact probability *P_c_*.
b. *C_b_* (aerosolizing) cases, who can generate airborne infectious particles that may transmit infection to others in shared air. These individuals also transmit via close contact, but additionally pose a risk of airborne transmission that is modulated by environmental conditions.

In each generation of transmission, new infections are generated through two routes:

a. Close-contact transmission, which follows Bernoulli trial logic consistent with Reed-Frost, with a probability of transmission:

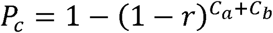

1. b. Airborne (aerosol) transmission, modeled using the Wells-Riley equation to generate an effective per-susceptible infection probability:

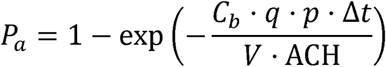

where *q* is the quanta generation rate (per aerosolizing individual per hour), *p* is the pulmonary ventilation rate (m³/hour), *V* is the volume of the shared airspace, ACH is the air changes per hour, and Δ*t* is the time step duration.

The joint probability of infection from either route is calculated as:

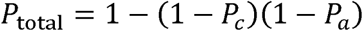

This simplifies simulation and ensures infections are not double-counted. However, it is also possible to use a mathematically equivalent de-overlapped formulation of the model, where close-contact infections are calculated first, and airborne transmission is applied to the remaining susceptibles not already infected via contact. This allows for route attribution but slightly underestimates overlap in risk (i.e., individuals infected by a contact transmitting *both* through close contact and aerosol infection).

The deterministic implementation of VAPOR assumes homogenous mixing of individuals and computes infection probabilities directly. In the stochastic VAPOR implementation, we introduce a dimensionless mixing parameter κ (assigned values between 0.05 and 0.15) to represent incomplete mixing and variability in spatial proximity. This parameter adjusts infection probabilities and avoids the unrealistic “all-or-none” outcomes of purely deterministic airborne transmission. In the stochastic formulation with mixing, close contact transmission probability is estimated as:

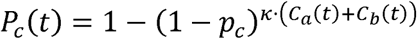

Airborne transmission probability is estimated as:

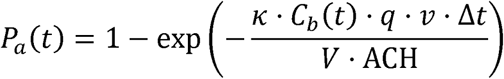

Joint probability of infection from either route is:

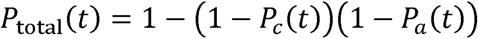

The number of new infections in each generation *t+1* is drawn from a binomial distribution:

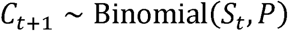

where *P* is either P_total_ or the combined outcome of sequential contact and aerosol draws (for route attribution), and *S_t_* is the number of susceptibles at time *t*. This stochastic formulation captures variability in transmission, the possibility of stochastic extinction, and a realistic distributions of outbreak sizes.

We assume a fixed fraction *f* of new cases are aerosolizing (*C_b_*), with the remainder *(1 - f)* in the *C_a_* group. In most simulations, we assume *f*=0.1, which matches Chen et al.’s empiric estimates for the earlier phase of the SARS-CoV-2 pandemic [cite Chen].

### iv. Estimation of R_0_ in Single-Patch Models

In the deterministic implementation of VAPOR, we derive a closed-form approximation of the basic reproduction number, *R*_0_, from its definition under the assumption of a fully susceptible population:

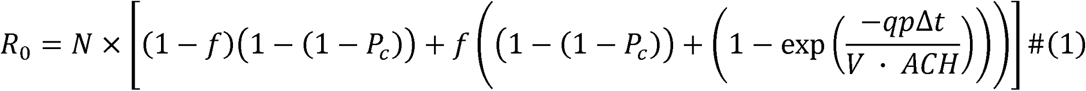

where *N* is the number of susceptibles at time zero, *P_c_*is the close-contact transmission probability per contact per generation, *q* is the quanta generation rate per person, *p* is the pulmonary ventilation rate of susceptibles, Δ*t* is the duration of the time step, and *v·ACH* is the clean air delivery rate, where ACH is air changes per hour and *V* is room volume. This closed-form expression captures the additive contribution of contact and aerosol-based transmission to the mean secondary case count.

In the stochastic implementation of VAPOR, transmission events are modeled as Bernoulli processes, with infections in each generation drawn from binomial distributions governed by the per-susceptible probability of infection. To estimate *R_0_*stochastically, we simulate multiple independent outbreaks (10,000 iterations) under identical initial conditions. For each simulation, the number of secondary infections generated in the first generation following seeding (generation 1) is recorded, and the reproduction number is estimated as:

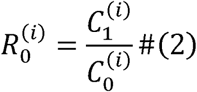

where *C_0_^(i)^* is the number of cases in generation 0 (initial seeding), and *C_1_^(i)^* is the number of new infections in generation 1 for the *i*^th^ simulation. Across all simulations, we compute the mean and distribution of *R*_0_. To characterize the degree of overdispersion in transmission, we fit a negative binomial distribution to the set of secondary case counts:

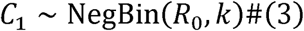

where *k* is the dispersion parameter, with lower values of *k* indicating greater heterogeneity and a higher probability of superspreading events. Fitting is performed using maximum likelihood estimation with the `fitdistr` function in R, and the resulting estimates of *R_0_* and *k* are used to quantify both the average and variability in transmissibility in the early stages of simulated outbreaks.

### v. Meta-population expansion of VAPOR

We extended VAPOR to model multiple interconnected spaces (“patches”), each characterized by distinct parameters including population size, ventilation rate (ACH), and air volume (*V*). Transmission within and between patches is governed by an exposure matrix, *E*, where each element *E_ij_* indicates relative exposure intensity from infectious individuals in patch *i* to susceptible individuals in patch *j*.

To address the issue of airborne transmission probabilities becoming overly deterministic (all or none), we introduced a mixing parameter κ (with typical values around 0.15 in our simulations), representing incomplete mixing of indoor air and variability in the spatial proximity between susceptible and infectious individuals. This parameter modulates exposure intensity, reducing the extremes of transmission probability and reflecting realistic spatial heterogeneity.

For a system of multiple patches indexed by *i* (infectious) and *j* (susceptible), the probabilities of infection are thus computed as follows:

For close-contact transmission,

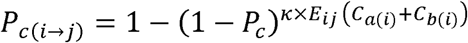

and for airborne (aerosol) transmission,

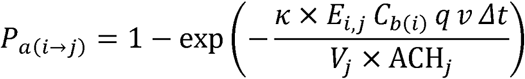

These probabilities are combined for each susceptible in patch *j*:

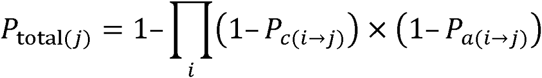

In each generation, the number of new infections in patch *j* is drawn from a binomial distribution:

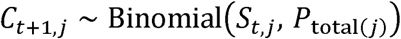

This formulation captures both route-specific transmission risks and the imperfect, heterogeneous mixing conditions typical of real-world indoor spaces.

### vi. Parameterization and Simulations

We simulated three-patch scenarios (patches A, B, and C) under various ventilation conditions, each scenario repeated 10,000 times to account for stochastic variability. Population size and air volume were identical across the three patches. The initial index case was seeded in patch A (“seed patch”) in all scenarios.

The ventilation scenarios included:

1. “All Low”: ACH = 0.5 in patches A, B, and C.
2. “Seed High”: ACH = 6 in the initial (“seed”) patch A, ACH = 0.5 in patches B and C.
3. “Other High”: ACH = 0.5 in the initial (“seed”) patch A and in patch B, ACH = 6 in patch C.
4. “Seed and Other High”: ACH = 6 in seed patch A and patch B, ACH = 0.5 in patch C.
5. “Others High”: ACH = 0.5 in seed patch A, ACH = 6 in patches B and C.
6. “All High”: ACH = 6 in patches A, B, and C.

These simulations allowed us to examine how ventilation in different patches impacted epidemic outcomes such as outbreak size. We performed sensitivity analyses in which we varied ventilation in “well ventilated” patches across a range from ACH = 2 to ACH = 12. We evaluated the independent contributions of ventilation in the seed patch, and good average ventilation across all patches, using negative binomial regression models.

The model was created using R version 4.5.0 (R Foundation for Statistical Computing, Vienna, Austria) run using R Studio version 2025.05.0 (Posit Software, Boston, MA). Model code is available at https://zenodo.org/badge/1026304551.svg)](https://doi.org/10.5281/zenodo.16422619.

## Results

### Deterministic Estimation of R_0_

We used the deterministic single-patch VAPOR model to estimate the basic reproduction number (*R*) across a range of ventilation conditions using Equation 1. As expected, increasing air changes per hour (ACH) led to substantial reductions in *R* due to decreased airborne risk, though the relationship was non-linear, with the largest relative reductions seen when ACH was low, and with diminishing returns at higher ventilation levels, irrespective of assumptions around quanta generation, fraction of aerosolizers, and room size (**Figure 1** and **Supplementary Figure S1**). The deterministic model also permits estimation of relative contributions to outbreak growth via contact and airborne routes over time, providing a platform for projection of impacts of source-specific intervention (e.g., handwashing vs. improved ventilation) (**Figure 1**).

**Figure 1.**
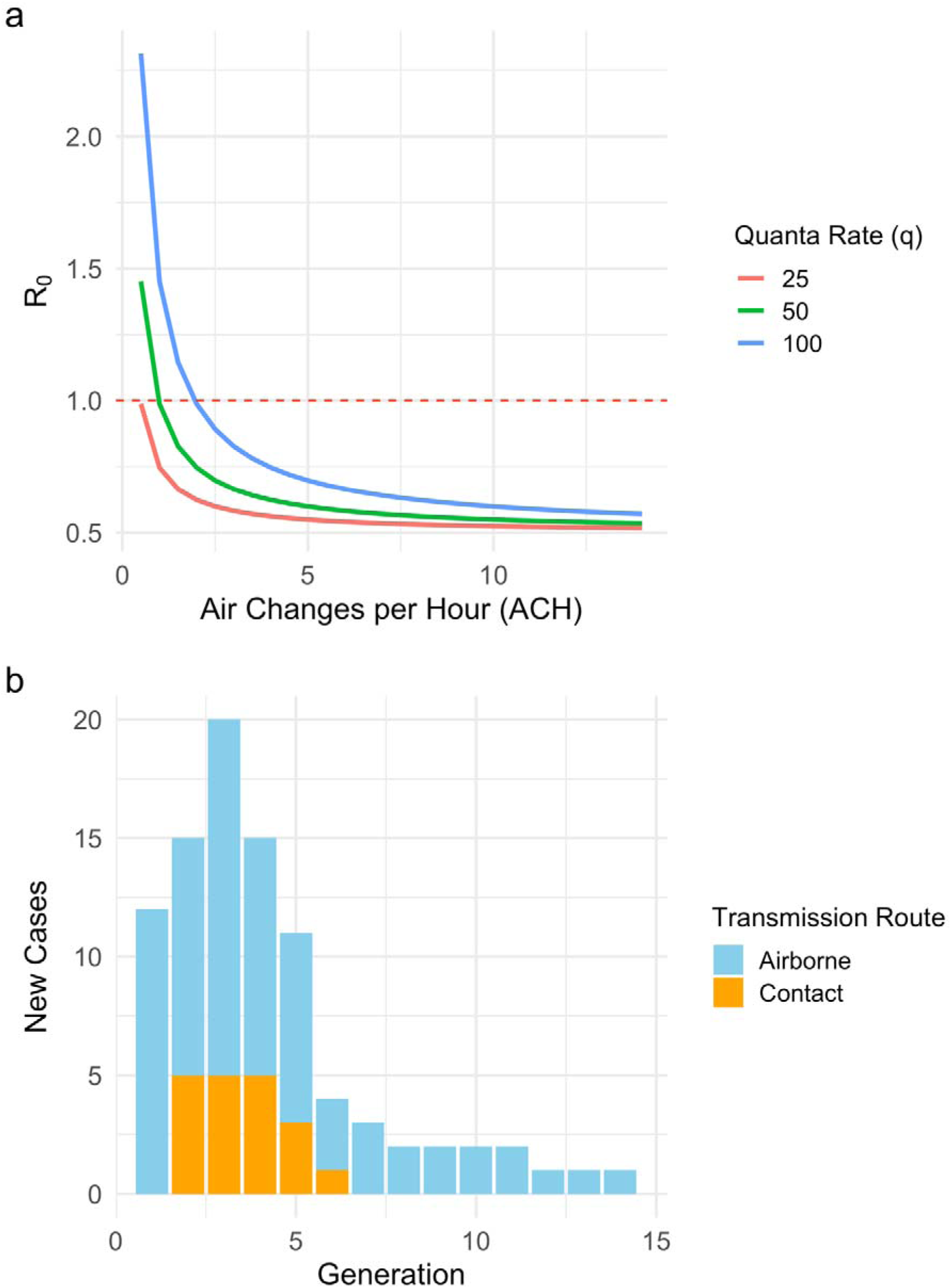
Impact of ventilation and transmission route on epidemic dynamics. Figure 1.a. (top), estimated basic reproduction number (R) as a function of air changes per hour (ACH), based on deterministic implementation of the VAPOR model. Curves are shown for three different aerosol quanta generation rates (q = 25, 50, 100), assuming 10% aerosolizing individuals. Figure 1.b. (bottom), de-overlapped attribution of new infections by route (contact vs. airborne) across epidemic generations under baseline conditions (ACH = 2, q = 50). Early transmission is dominated by airborne routes, emphasizing the disproportionate impact of poorly ventilated seed environments.

### Stochastic Estimation of R_0_

While the deterministic VAPOR model provides a tractable framework for calculating average transmission dynamics, it does not account for the random variation in transmission (e.g., stochastic extinction, super-spreading and long tails of outbreak size distributions) characteristic of emerging respiratory pathogens (18, 20, 21). To capture these critical real-world features, we developed a stochastic version of the VAPOR model. Across a range of quanta generation rates, ventilation levels (ACH), and proportions of aerosolizing index cases, there was close concordance between deterministic *R*_0_ values and average stochastic offspring counts, identical to *R*_0_ from Equation 2 with a single initial case (**Figure 2**). This confirms that the probabilistic structure of the stochastic model yields expected results under conditions where the law of large numbers holds, while also enabling quantification of variability and extinction that are not captured deterministically.

**Figure 2.**
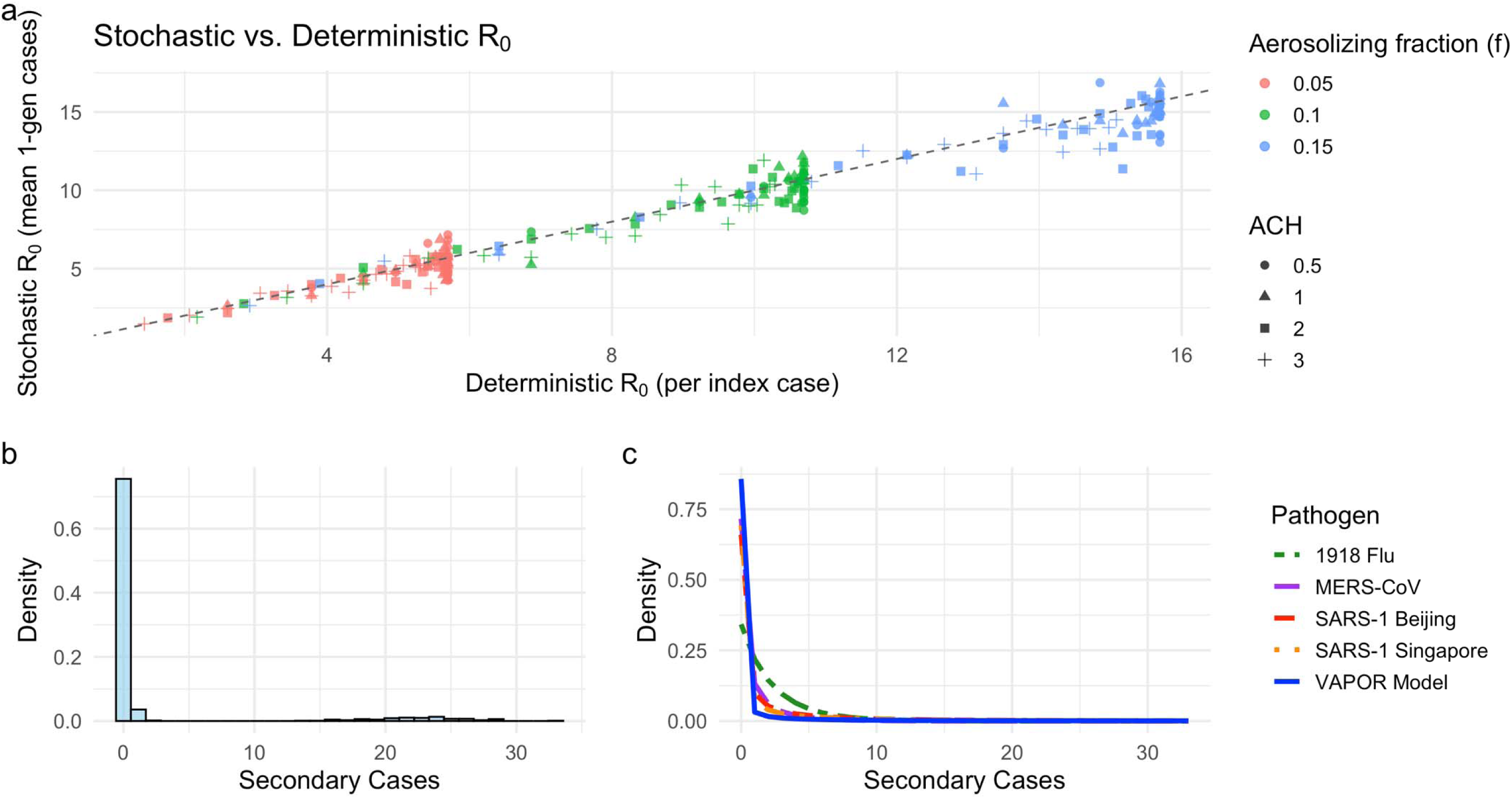
Stochastic realization of secondary transmission and offspring distributions. Figure 2.a. (top), comparison of deterministic and stochastic estimates of R using a single-generation model. Each point represents the mean number of secondary cases from 1,000 stochastic simulations at varying levels of ventilation (ACH), aerosolizing fraction *(f),* and quanta generation rate. The dashed line indicates parity between deterministic and stochastic expectations. Figure 2.b. (bottom left), histogram of secondary infections from simulated generation-1 outbreaks in a poorly ventilated space (ACH = 0.5, q=80q = 80q=80, f=0.15f = 0.15f=0.15). The distribution is strongly right-skewed, with most simulations producing zero or few secondary cases, and a minority producing many. Figure 2.c. (bottom right), negative binomial (NB) distributions comparing the VAPOR-derived offspring distribution to empirical estimates for 1918 influenza, MERS-CoV, and SARS-CoV-1 (Singapore and Beijing outbreaks). Like SARS-1 and MERS, the VAPOR model exhibits substantial overdispersion, consistent with superspreading dynamics in airborne-dominant transmission. The similarity highlights the role of environmental and stochastic factors in creating superspreading-dependence for respiratory pathogens such as emerging coronaviruses.

As initial simulations that assumed homogeneous risk in the population usually resulted in “all or none” outbreaks (entire population infected or no onward transmission) we modified our model by introducing a scaling parameter κ, which effectively serves as a proxy for incompletely mixed air. Using simulations of generation-1 offspring distributions, we estimated the average stochastic reproduction number (*R*) and dispersion parameter (*k*) by fitting a negative binomial distribution to the resulting secondary case data (Equation 3). Across 1,000 simulations under baseline conditions, we observed a mean *R* of 2.23 and a dispersion parameter *k* of 0.04, consistent with highly overdispersed transmission (**Figure 2**). We further conducted sensitivity analyses across a range of aerosol fractions (*f*), quanta generation rates (*q*), ventilation rates (ACH), and aerosol scaling (κ). The resulting estimates of *R* from 1st generation simulations are shown in **Figure 3**. As expected, lower ACH, lower κ, and higher quanta generation (infectivity per aerosolizing case) yielded greater transmission potential. The grid framework demonstrates that our stochastic implementation reproduces intuitive and epidemiologically plausible gradients in *R*, while incorporating stochastic extinction and variability absent from deterministic models.

**Figure 3.**
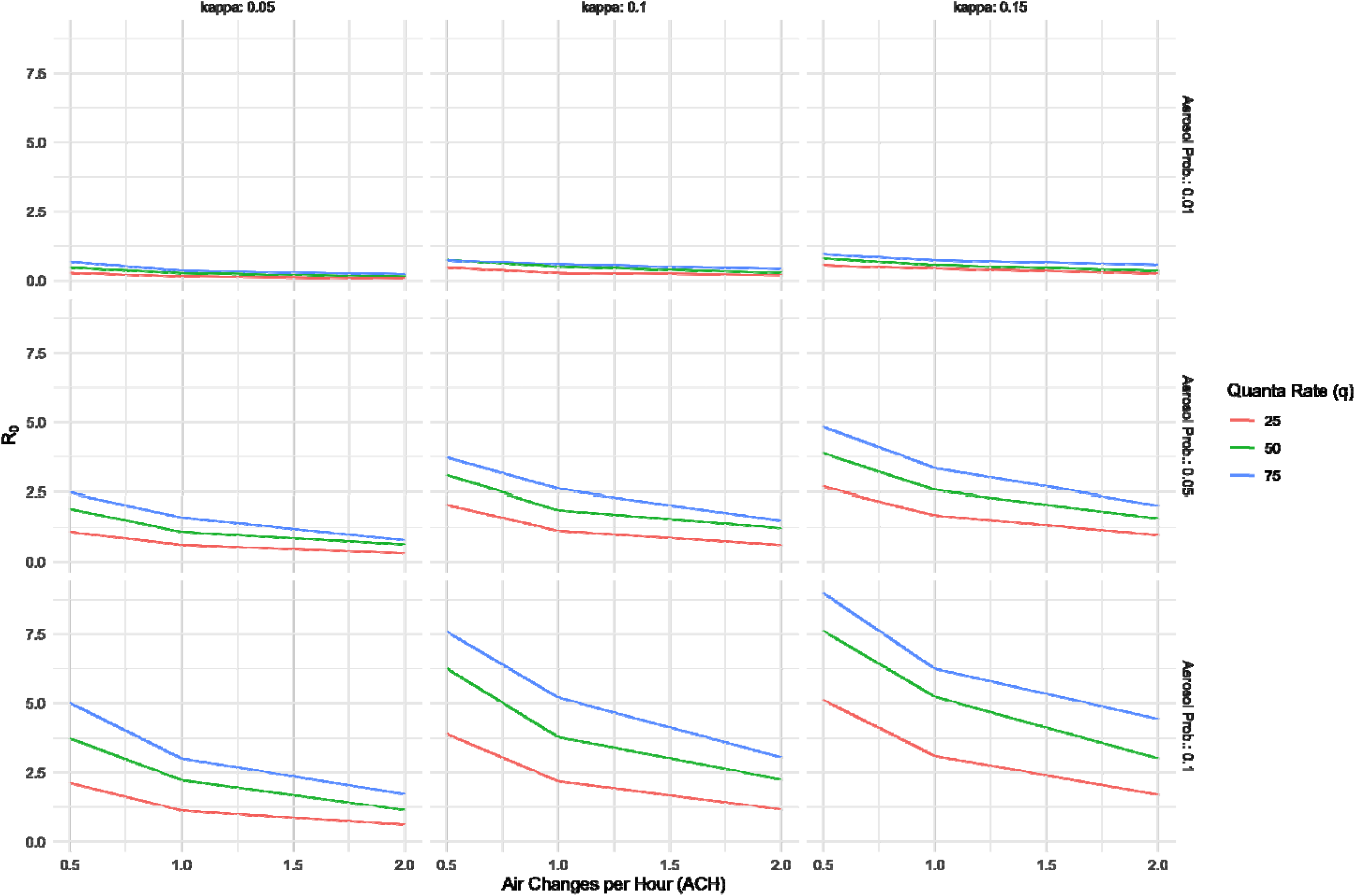
Sensitivity of stochastic R estimates to aerosol transmission parameters. We conducted a parameter sweep across aerosolizing fraction (f), quanta generation rate (q), ventilation rate (ACH), and the aerosol mixing parameter (κ). The resulting R estimates from first-generation stochastic simulations are displayed across panels (Y-axis). Lower ventilation rates (ACH; X-axis within panels), higher aerosolizing fractions (f; rows), and more uniform mixing (higher κ; columns) consistently yielded greater transmission potential. This grid illustrates how the VAPOR model produces intuitive and epidemiologically plausible gradients in R, while incorporating stochastic extinction and transmission variability absent from deterministic frameworks.

### Meta-population expansion of VAPOR

Simulations of the stochastic patch model confirmed a strong effect of ventilation, with both average ACH across the meta-population and high ACH in the “seed” patch where the index case emerged acting independently to significantly reduce case incidence, as evaluated with negative binomial regression (**Figure 4** and **Table 2**). There was no significant interaction between average ventilation and high ACH in the seed patch.

**Figure 4.**
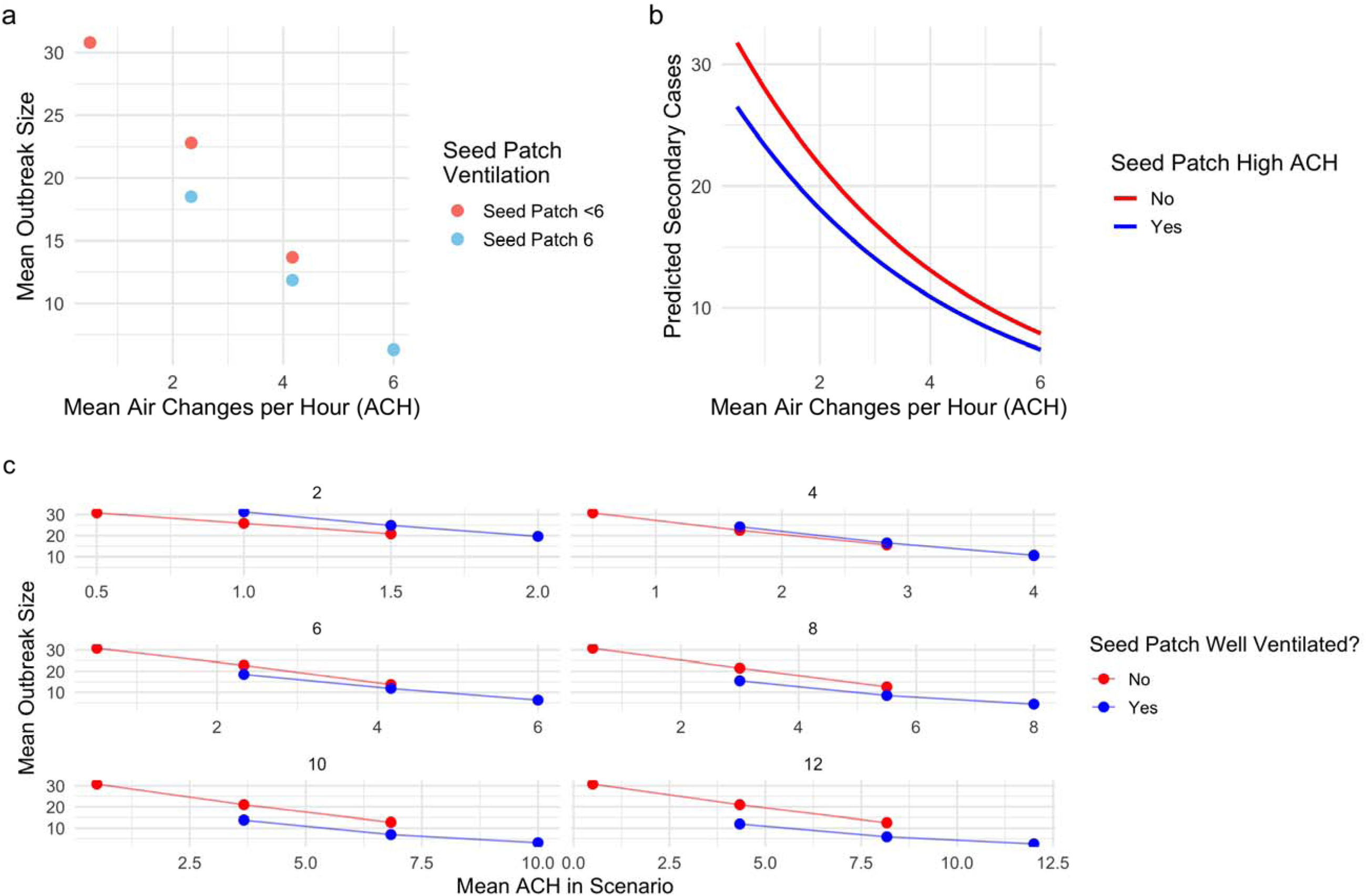
Effects of ventilation and seeding location on outbreak dynamics in a multi-patch stochastic transmission model. Figure 4.**a**. (top left), mean outbreak size (total infections across all patches) as a function of average air changes per hour (ACH) under six ventilation scenarios, stratified by whether the initial (seed) patch had high ventilation (ACH = 6; blue) or not (red). Figure 4.b. (top right), predicted number of secondary cases from a negative binomial regression model, showing a monotonic decline in outbreak size with increasing mean ACH, with an additional protective effect when the seed patch is well ventilated. Figure 4.c. (bottom), sensitivity analysis varying the threshold for defining “good ventilation” (x-axis facets show thresholds from 2 to 12 ACH). Each panel plots mean outbreak size by mean ACH for scenarios where the seed patch met the ventilation criterion (“Yes”, blue) versus not (“No”, red). A consistent divergence between the curves emerges only under more stringent ventilation thresholds (≥6 ACH), indicating that the benefit of high seed-patch ventilation is robust primarily when defining “good” ventilation as ≥6 ACH.

**Table 1.** Model Parameters.

| Parameter | Definition | Base Value | Source / Reference |
| --- | --- | --- | --- |
| $p_c$ | Close-contact probability of infection (per contact, per generation) | 0.00695 | Calibration, to achieve realistic $R_0$ estimates |
| $q$ | Quanta generation rate (quanta/hour/person) | 50 | (23-25) |
| $v$ | Pulmonary ventilation rate ($m^3$/hour) | 0.5 | (25, 34) |
| $\Delta t$ | Duration of exposure per generation (hours) | 96 | Approx. 4 days, reflecting Wuhan variant SARS-CoV-2 serial interval (35) |
| V | Volume of indoor airspace per patch (m<sup>3</sup>) | 500 | Representative classroom/office setting (29, 30, 36-38) |
| ACH (low) | Air changes per hour (baseline "low" ventilation scenario) | 0.5 | Low-end ventilation rate (29, 30, 36-38) |
| ACH (good) | Air changes per hour ("good" ventilation scenario) | 6 | CDC and ASHRAE recommendations for high ventilation (5-6 ACH) (29, 36-38) |
| $f$ | Fraction of cases aerosolizing (CbC_b cases) | 0.1 | (39) |
| $\kappa$ | Mixing parameter (dimensionless, stochastic model only) | 0.15 | Calibration, to avoid deterministic (all or none) aerosol transmission |
| $N$ | Initial number of susceptibles per patch | 100 | Initial disease emergence scenario ($R_0$ operative) |
| $C_0$ | Initial seed infectious individuals (index cases) per outbreak | 1 | Assumed single initial infection |

**Table 2.** Predictors of Outbreak Size from Negative Binomial Regression Model.

| Predictor | Incidence Rate Ratio | 95% CI | P-value |
| --- | --- | --- | --- |
| Mean ACH | 0.83 | 0.82 – 0.84 | < 0.001 |
| Seed Patch Well Ventilated (Yes vs No) | 0.48 | 0.43 – 0.53 | < 0.001 |

**NOTE:** Incidence rate ratios are exponentiated coefficients from a negative binomial regression model fitted to outbreak size (excluding the index case) in 10,000 simulations. A lower IRR indicates reduced outbreak size. “Mean ACH” corresponds to the mean air changes per hour across the environment. “Seed Patch Well Ventilated” indicates whether the patch where the infection started had high ventilation (ACH = 6 air changes per hour). Both higher mean ACH and initiating the outbreak in a well-ventilated patch were independently associated with significantly smaller outbreaks.

By contrast, in simulations of this overdispersed communicable disease with a high baseline probability of stochastic extinction, we found no robust increases in the probability of stochastic extinction (whether defined as <1, <5, or <10 secondary cases) with improved ventilation (see **Supplementary Appendix**). Sensitivity analyses in which the definition of “good” ventilation was varied from 2 to 12 ACH identified an inverse dose–response relationship between ventilation in the seed patch and outbreak size, but also diminishing returns with improvements in average ACH across the meta-population once 6 ACH was exceeded (**Figure 4**). No stable effects of improved ventilation on the probability of stochastic extinction were identified in these sensitivity analyses.

## Discussion

We created a hybrid modeling approach that integrates a canonical cross-sectional airborne risk model from engineering with a canonical dynamic outbreak model from epidemiology, to better understand, manage, and mitigate the emergence of airborne respiratory diseases and outbreaks (5, 6, 26–28). By combining the Wells-Riley model (8), which explicitly quantifies infection risk as a function of ventilation and air quality, with the generational structure of the Reed-Frost outbreak framework (14), the VAPOR model provides a flexible platform for simulating the role of both shared environments and direct contact in the emergence and spread of airborne pathogens. The ease with which these two canonical frameworks can be combined underscores the conceptual compatibility of engineering and epidemiological approaches, despite differences in emphasis and vocabulary.

This hybrid approach allowed us to show that outbreaks seeded in poorly ventilated environments contribute disproportionately to disease emergence and transmission. The worst-ventilated areas pose the greatest risk if they happen to be the location where a novel disease emerges. In other words, the infection control chain is only as strong as its weakest link. The model also permits route-specific analysis, making it possible to distinguish between close-contact and airborne transmission, and provides a platform to explore the impact of different types of intervention. Our realization of the model conceptualized clean air delivery as a function of ventilation, though newer guidance from organizations such as ASHRAE emphasizes the use of “clean air delivery rates” (29), which can be achieved by ventilation, or by filtration and decontamination with inexpensive and scalable tools (e.g., portable filters and/or high room UV light sources). The Wells-Riley equation can be readily adapted to incorporate such additional approaches (12), and consequently so can our model.

We found that the model could be realized in either deterministic or stochastic form, with strong concordance in *R* estimation regardless of the approach. However, our results highlight that stochastic simulations are essential for capturing the reality of early outbreak phases under the overdispersed transmission patterns characteristic of recent emergences, where random extinction or superspreading can strongly influence epidemic trajectories (17, 18, 20, 21).

Importantly, we found that for a highly overdispersed communicable disease like that simulated here, the impact of ventilation on rapid reduction in reproduction numbers is easier to demonstrate than is the impact on probability of emergence, as most introductions (regardless of ventilation) fail to result in secondary propagation. Nonetheless, we demonstrate a clear role for ventilation improvements in containing emergences more rapidly, which in the real world should translate into prevention of sustained epidemics and pandemics.

Recent developments in indoor air quality standards, such as the NIOSH recommendation of at least 5 air changes per hour (30) and the adoption of ASHRAE Standard 241-2023 (29), mark a fundamental shift in infection control policy, recognizing that ventilation and air cleaning are now core components of epidemic risk management in all occupied indoor spaces, not just healthcare settings. ASHRAE 241-2023 is the first comprehensive building standard focused on infectious aerosol control and established minimum requirements for equivalent clean airflow that are tailored to occupancy and space type, allowing building operators the flexibility to meet these targets through any combination of ventilation, filtration, or air cleaning.

These new standards reflect a growing scientific consensus that the risk of airborne transmission is best mitigated not by marginal improvements to average ventilation, but by prioritizing the most poorly ventilated or highest-risk (i.e., highest probability of being the site of an initial seed case) environments, a finding directly supported by our modeling results. The VAPOR framework demonstrates the disproportionate effect that interventions in these “weakest link” settings can have on epidemic dynamics and illustrates how achieving or exceeding the new recommended airflow standards can substantially reduce outbreak potential. While the VAPOR model used ACH, other formulations of the Wells-Riley equation can incorporate other interventions such as portable filter, germicidal UV, or mask and respirator use rather than air replacement (29). The convergence of scientific evidence, engineering standards, and public health policy underscores the urgency and attainability of making clean indoor air a foundation of epidemic and pandemic preparedness and equitable infection control. It is also worth noting that clean indoor air can act synergistically with other public health measures, like vaccination (31), to mitigate and prevent outbreaks, and this interaction will be our focus in future work. A large and growing body of health economic literature demonstrates that while the costs of improving indoor air quality are substantial, they are dwarfed by the costs of disease and disruption that results from airborne infectious and non-infectious diseases (4). Bruns has suggested that the benefit to cost ratio of widespread implementation of ASHRAE 241-2023 and ASHRAE 62.1 would be between 10:1 and 40:1 (32, 33).

Our study has several limitations that should be acknowledged. First, while the VAPOR model captures key features of airborne transmission and ventilation heterogeneity, it remains a simplification of real-world complexity. We did not account for dynamic behavioral changes in response to outbreaks, nor for heterogeneity in host susceptibility or pathogen characteristics that may evolve during an epidemic. The meta-population structure, while valuable for illustrating heterogeneity across linked spaces, assumes relatively simple movement and mixing patterns, and does not yet incorporate networked or gravity-based connectivity, which are likely to be especially relevant in larger or urban systems. Parameter choices for quanta generation, mixing, and fraction of aerosolizers, while empirically informed, remain subject to uncertainty and may vary by pathogen, activity, and setting. Moreover, while the Wells-Riley component of our model can incorporate other interventions such as filtration, UV disinfection, or masking (12), we did not explicitly simulate the combined effects of these interventions alongside ventilation in this initial analysis.

Future work should address these limitations in several ways. A natural next step is the extension of VAPOR to incorporate gravity model structures, allowing for more realistic representation of movement and transmission between heterogeneous environments, such as between schools, workplaces, and homes within a community. Incorporation of vaccination dynamics, both for individual-level protection and population-level herd effects, will also be essential for exploring synergies between air quality interventions and immunization programs. Most importantly, future modeling efforts should seek calibration and validation against real-world outbreak data from diverse settings and pathogens. This will allow for refinement of model parameters, testing of intervention strategies, and greater confidence in policy recommendations. By building on the flexible and transparent structure of VAPOR, these advances will help bridge the gap between engineering, epidemiology, and public health implementation, ultimately supporting more effective and equitable control of airborne infectious diseases. As improved indoor air quality standards become more widely adopted, hybrid modeling approaches like VAPOR will be increasingly important for informing practical and equitable strategies to reduce the burden of airborne infectious diseases.

## Data Availability

Model code is available at https://zenodo.org/records/16422619. Further guidance related to the model can be obtained by contacting Dr. Fisman.

## Supplementary Appendix

**Supplementary Figure S1.**
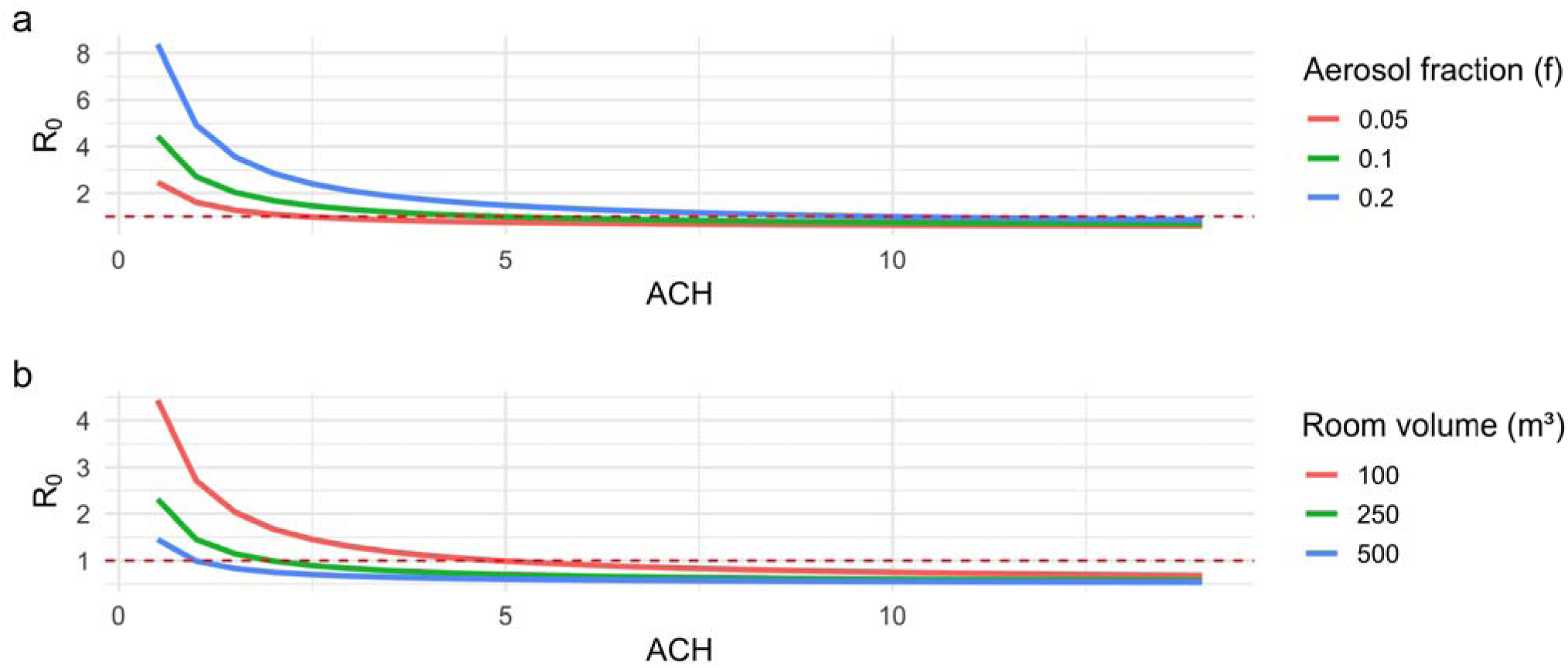
Sensitivity of R to ventilation, room volume, and aerosol-generating fraction. Panel (a), estimated R as a function of air changes per hour (ACH) across varying aerosolizing fractions (*f* = 0.05, 0.1, 0.2). Higher aerosol fractions yield greater transmission potential, particularly at lower ACH values, with diminishing impact as ventilation improves. Panel (b), **e**stimated R as a function of ACH under different room volumes (100 m³, 250 m³, 500 m³). Larger room volumes reduce airborne transmission risk by diluting quanta concentration, resulting in lower R across all ACH values. Dashed horizontal line at R = 1 denotes the epidemic threshold.

**Supplementary Figure S2.**
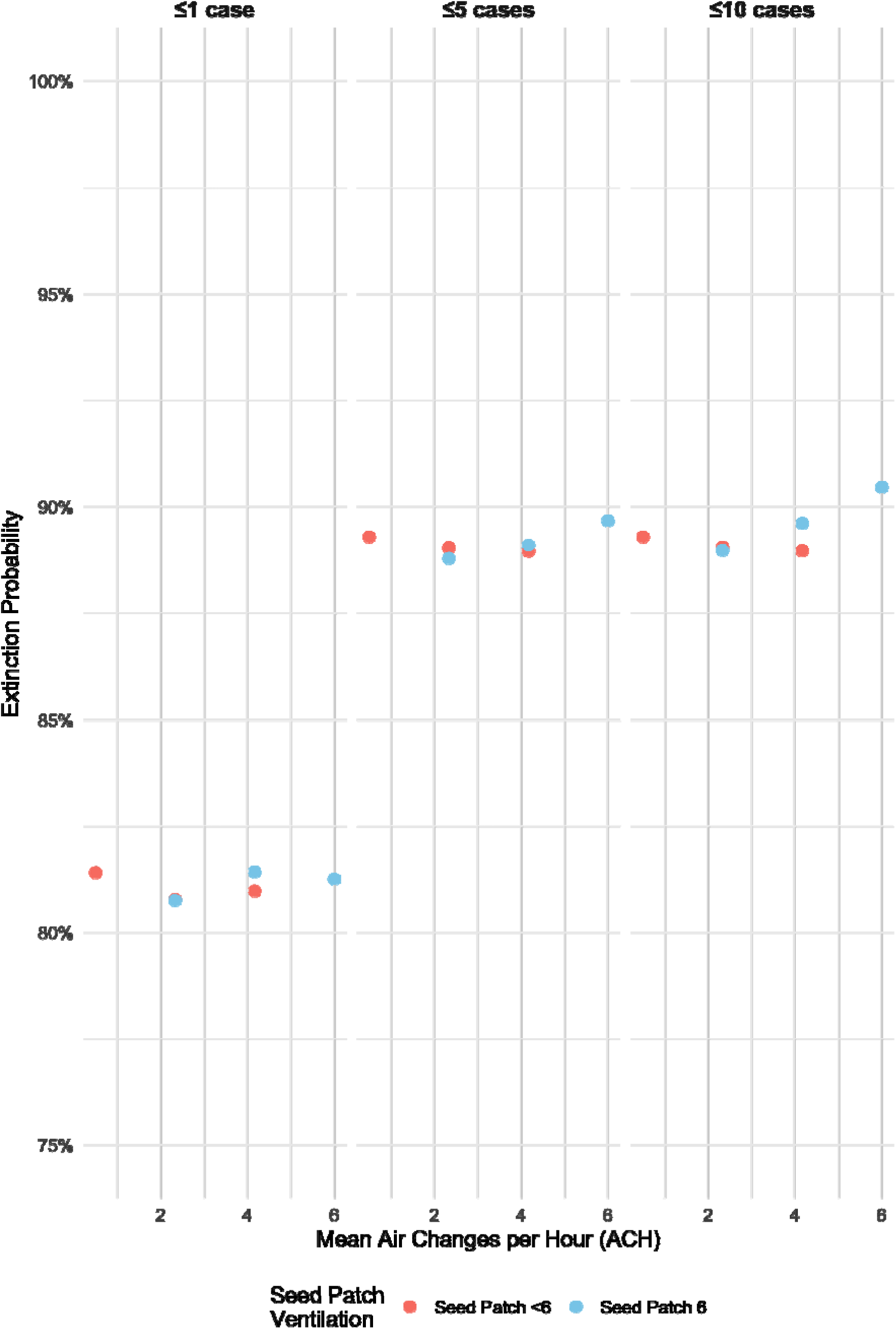
Extinction probabilities across ventilation scenarios and exctinction thresholds. Probability of outbreak extinction is shown as a function of mean air changes per hour (ACH) in the environment and whether the seed patch was well-ventilated (ACH = 6). Results are stratified by extinction threshold, defined as total outbreak size ≤1 case, ≤5 cases, or ≤10 cases. In contrast to outbreak size, which decreased with increasing mean ACH and good ventilation in the seed patch, there was no clear increase in the probability of exctinction with increasing ACH or good seed patch ventilation. For the markedly overdispersed disease that we simulated, extinction probability is high in all scenarios. Data represent 10,000 stochastic simulations per scenario.

---

**Supplementary Table S2.** Logistic Regression Odds Ratios for Extinction Extinction odds ratios for thresholds of ≤1 case, ≤5 cases, and ≤10 cases. Odds ratios are exponentiated coefficients from logistic regression models using mean air changes per hour (ACH) and whether the seed patch was well-ventilated (ACH = 6) as predictors. There is no consistent association between ventilation and the probability of extinction. Data reflect 10,000 stochastic simulations per scenario.

**Threshold: Extinction = $\leq 1$ case**
| Predictor | Odds Ratio | 95% CI | p-value |
| --- | --- | --- | --- |
| Mean ACH | 0.9886 | (0.9759, 1.0016) | 0.097 |
| Seed Patch 6 ACH | 1.0029 | (0.9553, 1.0531) | 0.905 |

**Threshold: Extinction = $\leq 5$ cases**
| Predictor | Odds Ratio | 95% CI | p-value |
| --- | --- | --- | --- |
| Mean ACH | 0.9935 | (0.9765, 1.0108) | 0.454 |
| Seed Patch 6 ACH | 1.0155 | (0.9563, 1.0783) | 0.615 |

**Threshold: Extinction = $\leq 10$ cases**
| Predictor | Odds Ratio | 95% CI | p-value |
| --- | --- | --- | --- |
| Mean ACH | 1.002 | (0.9850, 1.0193) | 0.821 |
| Seed Patch 6 ACH | 1.059 | (0.9970, 1.1254) | 0.063 |

